# Measuring health system quality with routine health information systems in Rwanda

**DOI:** 10.1101/2024.10.24.24316072

**Authors:** Celestin Hategeka

**Affiliations:** Centre for Health Services and Policy Research, School of Population and Public Health, Faculty of Medicine, University of British Columbia, Vancouver, BC, Canada

**Keywords:** Health system quality, Quality of care, quality measurement, routine health information systems

## Abstract

**Background:** Growing evidence suggests that achieving the Sustainable Development Goal (SDG) 3 will require high-quality health systems in low and middle-income countries. The objective of this study was to assess whether routine health information systems in Rwanda capture relevant health system quality measurements to facilitate the effective tracking of the Rwandan health system performance.

**Methods:** I systematically reviewed the Rwanda health management information systems (Rwanda HMIS)—one of the six core building blocks of health systems—to identify health system performance indicators corresponding to processes of care quality and quality impact dimensions of high-quality health systems proposed by the Lancet Global Health Commission on High-Quality Health Systems in the SDG Era. Using a cross-sectional study design and descriptive statistics, I summarized available quality indicators by domains of the high-quality health system framework.

**Results:** Overall, less than 30% of the indicators collected in the Rwanda HMIS are processes of care quality and / or quality impact indicators. Health outcome measures were captured across health center and hospital HMIS reporting forms. However, there were gaps in the measurement of relevant quality impact measures such as confidence in health systems and economic benefit, and processes of care quality measures such as user (patient) experience, safety, continuity, and integration of care. Measurements about competent care and systems care were rarely available outside maternal, newborn, and child health.

**Conclusion:** The current routine health information systems in Rwanda would benefit from capturing additional healthcare quality metrics, including for noncommunicable diseases, to allow the effective tracking of the health system performance and to identify new potential efficiencies to maximize the impact of the Rwandan health system.

## Introduction

Growing evidence suggests that achieving Sustainable Development Goal (SDG) 3 will require not only universal health coverage but also high-quality health systems in low- and middle-income countries (LMICs) [1–3]. The Lancet Global Health Commission on High-Quality Health Systems in the SDG Era recently identified measuring and reporting health system performance metrics that matter most to people such as processes of care quality and quality impacts as a key to accountability and improvement of health systems [1]. Although processes of care and quality impacts more accurately reflect the return on investments in the health system and its potential to improve health outcomes, information on these metrics remain scarce in many LMICs [4]. For example, Macarayan et al (2018) assessed the quality of primary care using Service Provision Assessment (SPA) surveys—the most comprehensive and nationally representative surveys of health systems in many LMICs—and found that these surveys capture limited information on key elements on the quality of primary care. Similarly, Demographic and Health Surveys (DHS) conducted in many LMICs do not capture many key elements of health system quality [1].

While surveys such as SPA and DHS have advantages of being consistent and thus allowing comparability across countries, they are done sporadically and as such they do not provide timely and longitudinal information on the performance of health systems. For example, the most recent SPA survey conducted in Rwanda was completed in 2007 and is thus likely not reflective of more recent performance of the Rwandan health system. Routine health information systems (RHIS) or health management information systems (HMIS), one of the six core building blocks of health systems, have been rapidly expanding in Rwanda like in many other LMICs over the last several years. They are currently operational in 67 LMICs with 2.28 billion population or 30% of the world’s population and are a primary source of health system performance [5–7]. They offer distinct advantages, including timely and longitudinal population-based information, making it possible to effectively track the performance of health systems to identify new potential efficiencies in a timely manner.

However, routine health information system data have not widely been used to study health system performance. Similarly, the extent to which relevant aspects of health system performance that matter most to people are captured in RHIS/HMIS is not well known in Rwanda. Evidence from other settings including Ethiopia, Kenya, Nepal, and Senegal suggests that the availability of processes of care quality and quality impact measures in routine health information systems vary across settings, but the overall capture of these indicators is limited [1]. Overall, RHIS in many LMICs have largely focused on collecting data on inputs or service volume [1]. While inputs should be measured as they are foundational to healthcare provision, they offer a limited insight into health system quality [1]. Existing evidence has shown weak relationships between input metrics and processes of care quality, highlighting a need to capture and track processes of care quality and quality impacts [1].

The objective of this study was to assess whether RHIS in Rwanda capture relevant health system quality measures that can be used to track the performance of the health system to identify new potential efficiencies to inform strategies to maximize the system impact.

## Methods

### Study design ad Data source

This study used a cross-sectional study design to describe the availability of quality indicators in the Rwanda health management information system (Rwanda HMIS). I reviewed the Rwanda HMIS reporting forms available from the Rwanda Ministry of Health website to assess whether relevant health system quality measures are captured [8]. The Rwanda HMIS is the routine health information system in Rwanda which captures health information including on care seeking and volume of services provided across Rwandan health facilities [9]. Data are aggregated at facility level and reported each month. The Rwanda HMIS was upgraded to DHIS2 in 2012, which changed the data collection methods [9, 10]. All indicators captured in the most recent versions of Rwanda HMIS Health Center reporting form and Rwanda HMIS Hospital reporting form were abstracted and entered into an Excel database verbatim.

### Choice of the framework for analysis

I used the framework of the Lancet Global Health Commission on High-Quality Health Systems in the SDG Era to classify indicators as quality indicators or otherwise [1]. This framework was chosen because it expands on previous frameworks, including the Donabedian framework widely used for evaluation of quality of care, by providing detailed attributes that can be measured to assess the performance of health systems from a quality of care lens. Specifically, it consists of three domains: foundations, processes of care, and quality impacts [1]. Foundations include the population (and their health needs and expectations), governance of health sector, platforms for care delivery, health workforce, and tools or resources [1]. Processes of care consists of competent care and systems, and positive user experience, while quality impacts include health outcomes, confidence in system and economic benefit and equity [1]. The framework emphases on having the capacity to measure and use data for learning and continuous improvement [1].

Quality indicators that were considered for inclusion in this study corresponded to two of the three dimensions of the Commission framework (processes of care and quality impacts) as summarized in Table 1 [1]. The Rwanda HMIS was also reviewed for equity stratification factors such as socioeconomic status (wealth index and education) and area of residence (rural vs urban) relevant to the demographics dimension, one of the three dimensions of vulnerability to poor quality of care highlighted by the Commission [1].

**Table 1.** Components of the two dimensions of the framework of the Lancet Global Health Commission on High-Quality Health Systems in the SDG Era.

| **Domains of processes of care** | **Sub-domains of processes of care** |
| --- | --- |
| Competent care or evidence-based, effective care | Systematic assessment, correct diagnosis, appropriate treatment, counselling, and referral. |
| Competent systems | Safety, prevention and detection, continuity and integration, timely action |
| Positive user experience | Respect: dignity, privacy, non-discrimination, autonomy, confidentiality, and clear communication; and user focus: choice of provider, wait times, patient voice and values, affordability, and ease of use |
| **Domains of quality impacts** | **Sub-domains of quality impacts** |
| Health outcomes | Morbidity, facility mortality, stillbirth, quality of life |
| Confidence in system | Satisfaction, recommendation, trust, and care uptake and retention |
| Economic benefit | Work production loss, reduction in health system waste, and financial risk protection |

SDG, Sustainable Development Goal.

### Analysis

I performed descriptive analysis using frequencies (and related percentages) to summarize available quality indicators by domains of the framework of the Lancet Global Health Commission on High-Quality Health Systems in the SDG Era. Additionally, I classified indicators as patient-reported if the data were collected using patient self-report. Although most indicators were similar across health center and hospital reporting forms, there were some differences because of different type of services provided at various levels of care in Rwanda. As such, I stratified the analysis by type of Rwanda HMIS reporting forms (health center and hospital) and further by maternal newborn and child health (MNCH) and other services.

## Results

Table 2 and Figure 1 summarize quality indicators available in the Rwanda HMIS. As shown in the Figure, more than 70% of the indicators collected in the Rwanda HMIS at the health center and hospital level are not processes of care quality or quality impact indicators. I found that many health outcome measures were captured across health center and hospital HMIS reporting forms. However, there were gaps in the measurement of relevant quality impact measures such as confidence in health systems and economic benefit, and processes of care quality measures such as user (patient) experience, safety, continuity and integration of care. Appendix tables 1 and 2 provide examples of indicators currently captured in the Rwanda HMIS.

**Figure 1.**
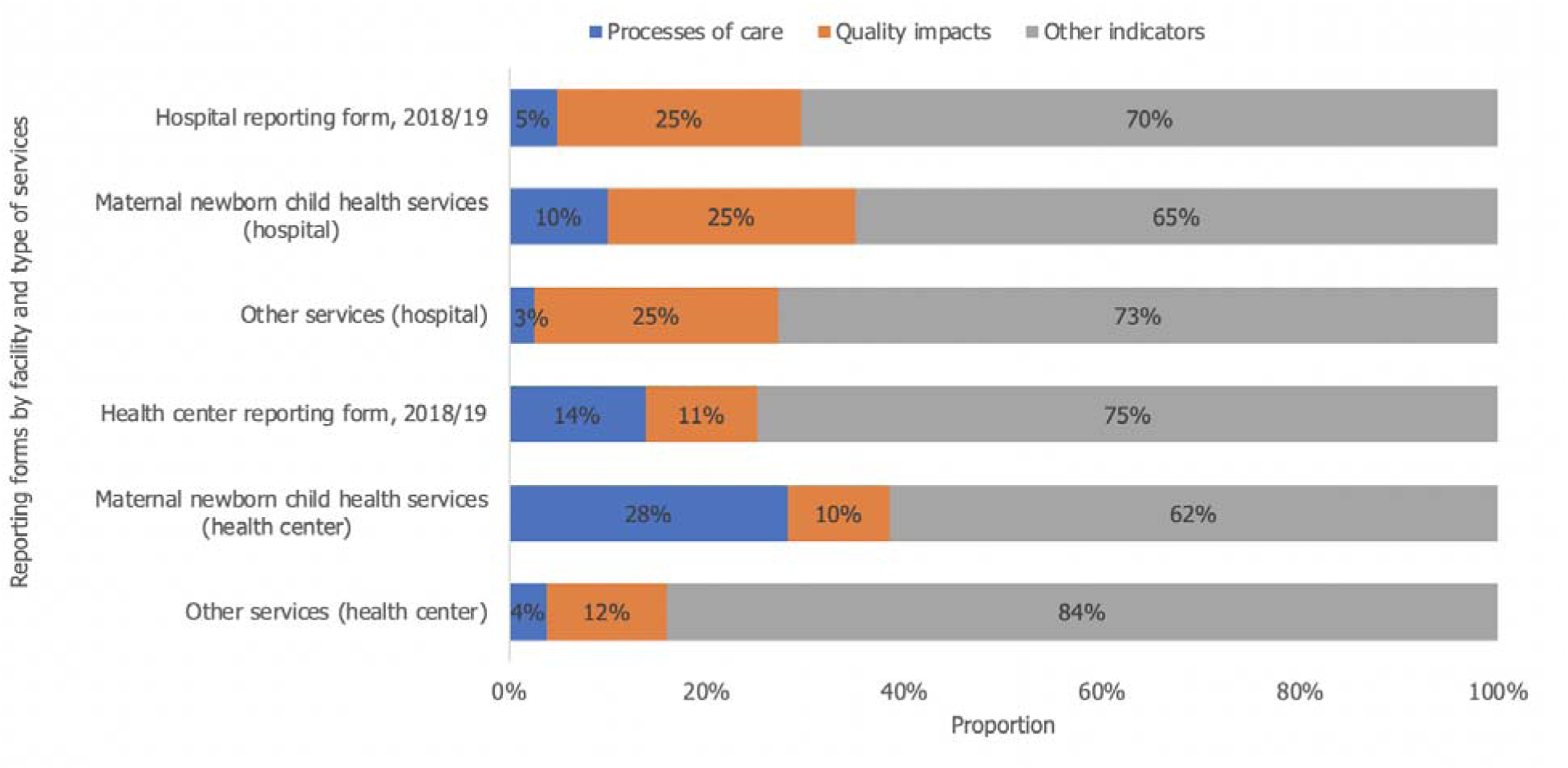
Proportion of Rwanda health management information system indicators by the two dimensions of the high-quality health systems framework or otherwise. Other indicators generally included volume of services provided and input indicators.

**Table 0.** Number of indicators from the Rwanda HMIS mapped into the high-quality health systems framework.

|  | Indicators | Processes of care |  |  | Quality impacts |  |  | Patient-reported |
| --- | --- | --- | --- | --- | --- | --- | --- | --- |
|  |  | Competent care<sup>†</sup> | Competent system<sup>‡</sup> | User experience | Health outcomes<sup>¶</sup> | Confidence | Economic benefit |  |
| HMIS reporting forms, n |  |  |  |  |  |  |  |  |
| Health center reporting form, 2018/19 | 573 | 52 | 27 | 0 | 65 | 0 | 0 | 0 |
| • MNCH | 234 | 44 | 22 | 0 | 24 | 0 | 0 | 0 |
| • Others | 339 | 8 | 5 | 0 | 41 | 0 | 0 | 0 |
| Hospital reporting form, 2018/19 | 727 | 28 | 7 | 0 | 180 | 0 | 0 | 0 |
| • MNCH | 220 | 20 | 2 | 0 | 55 | 0 | 0 | 0 |
| • Others | 507 | 8 | 5 | 0 | 125 | 0 | 0 | 0 |

Indicators were mapped against the processes of care and quality impact domains of the high-quality health systems framework, identifying the single most relevant for each indicator. Moreover, I classified indicators as patient-reported if data patient reported. Cells are colored by the availability of relevant indicators per row, with orange indicating no information available and blue (some) information available for that measurement set.
<sup>†</sup>Competent care sub-domain captured in the Rwanda HMIS (assessment, appropriate treatment, counselling, and referral); <sup>‡</sup>competent system sub-domain captured in the Rwanda HMIS (prevention / detection, timely care); and <sup>¶</sup>health outcome sub-domain captured in the Rwanda HMIS (mortality, stillbirth, morbidity). MNCH, maternal newborn and child health; HMIS, health management information system.

Measurements about competent care and systems and outcomes such as morbidity due to poor quality care were rarely available outside maternal, newborn, and child health. Many of the available processes of care indicators were added into the Rwanda HMIS reporting in 2018 (Table 3). For example, of the 573 indicators in health center reporting form, 116 (20.2%) were newly added, of which 11 (9.5%) were processes of care indicators and 18 (15.5%) were quality impact indicators. Similarly, of 727 indicators in hospital reporting form, 162 (22.3%) were newly added indicators, of which 7 (4.3%) were processes of care and 43 (26.5%) quality impacts. The newly added processes of care quality metrics include maternal and newborn health indicators such as antenatal corticosteroid therapy for risk of premature delivery, oxytocin for active management of third stage of labor (AMTSL), and antibiotics for preterm premature rupture of the membranes (PPROM).

**Table 3.** Number of indicators that were newly added to the Rwanda HMIS.

|  | All newly added indicators | Processes of care | Quality impacts | Other indicators |
| --- | --- | --- | --- | --- |
| Health center reporting form, 2018/19, n | 116 | 11 | 18 | 87 |
| MNCH services, HC | 41 | 10 | 6 | 25 |
| Other services, HC | 75 | 1 | 12 | 62 |
| Hospital reporting form, 2018/19, n | 162 | 7 | 43 | 112 |
| MNCH services, Hospital | 57 | 6 | 14 | 37 |
| Other services, Hospital | 105 | 1 | 29 | 75 |

HMIS, health management information system; MNCH, maternal, newborn, and child health; HC, health center; Other indicators generally included volume of services provided and input indicators.

Indicators for MNCH included antenatal care (ANC), childbirth, and postnatal care (PNC). Information on the WHO’s guidelines for Basic Emergency Obstetric and Newborn Care (BEmONC) interventions provided at the primary level of care were also captured in Rwanda HMIS: Intravenous antibiotics to manage obstetrical infections; Mother received parenteral uterotonic drugs (oxytocin) to manage PPH; Manual removal of placenta; Post-abortion care (manual vacuum aspiration or curettage to remove retained products of conception); Delivery by vacuum extraction; and (Pre) eclampsia cases receiving magnesium sulfate. Similarly, information on the WHO’s guidelines for

Comprehensive Emergency Obstetric and Newborn Care (CEmONC) interventions provided at the secondary and tertiary levels of care were captured in the Rwanda HMIS: CEmOC interventions include the signal functions of BEmOC listed above, plus surgery (e.g. caesarean section) and blood transfusion for obstetrical complications.

Measures on dimensions of vulnerability to poor quality of care such as socioeconomic status highlighted by the Lancet Global Health Commission on High-Quality Health Systems in the SDG Era were not available, making it difficult to conduct equity analysis using the Rwanda HMIS data. However, age and sex stratifiers are captured in the Rwanda HMIS.

## Discussion

The objective of this study was to assess whether routine health information systems in Rwanda capture relevant health system quality measures. I found that less than 30% of the indicators collected in the Rwanda HMIS are processes of care quality and / or quality impact indicators. Health outcome measures were captured across health center and hospital HMIS reporting forms. However, there were gaps in the measurement of relevant quality impact measures such as confidence in health systems and economic benefit, and processes of care quality measures such as user (patient) experience, safety, continuity, and integration of care. Measurements about competent care and systems care were rarely available outside maternal, newborn, and child health.

Research suggests that processes of care and quality impact indicators more accurately reflect the returns on investments in the health systems and their potential to improve health outcomes; however, information on these indicators appears limited in Rwanda echoing from findings the Lancet Global Health Commission on High-Quality Health Systems in the SDG Era [4]. For example, Kruk and colleagues found that, of the 121 indicators captured in routine health information systems in Ethiopia (Ethiopian HMIS) in 2014, only approximately 11 (9%) were processes of care indicators and 11 (9%) quality impact indicators [4]. Similar to the Rwanda HMIS, the Ethiopian HMIS did not capture any quality indicators relevant to user experience or confidence in health system or economic benefit; and none of the indicators were patient-reported, although this is integral to achieving patient-centered health systems [4].

A review of the Kenya’s routine health information system (Kenya HIS) showed that in 2012 it captured 198 indicators, of which about 56 (28.3%) were process of care quality indicators and 17 (8.6%) quality impact indicators [4]. Unlike the Rwanda HMIS, the Kenya HIS also captured some information on user experience and some of the indicators were patient-reported [4]. Routine health information systems in Senegal capture data on indicators such as user experience and people confidence in health systems [4]. Josephson and colleagues analyzed 68 quality checklists from 28 LMICs to understand how performance-based financing programs measure quality of care and they found that only 19% of indictors were processes of care indicators, 1% were quality impact (outcome) indicators, and 80% were structure indicators [11].

Many of the available health system quality measures appear to have been captured in the Rwanda HMIS from 2018 and onward, suggesting that Rwanda has been making efforts to increase the number of quality measures in its routine health information systems. The newly added processes of care quality metrics such as antenatal corticosteroid therapy for risk of premature delivery, oxytocin for AMTSL and antibiotics for PPROM will aid in the effective tracking of the Rwandan health system performance and can inform strategies to improve care competence. Some of these newly added indicators (e.g., antenatal corticosteroid therapy, oxytocin for AMTSL and antibiotics for PPROM) correspond to clinical practice guidelines that have been disseminated to health providers through the Advanced Life Support in Obstetrics (ALSO) or Emergency Obstetrical and Neonatal Care (EmONC) training program in Rwanda. As such, tracking longitudinal changes in these indicators can help in evaluating the uptake of ALSO clinical practice guidelines. Increasingly, data-driven quality improvements have been advocated to contribute to improving quality of care in LMICs [12, 13]. Judicious addition of process of care quality indictors related to other quality improvement programs (e.g., IMCI and ETAT+) would also go a long way to help in the evaluation of uptake of these programs over time. Similarly, adding equity metrics such as socioeconomic status could help identify who is getting lower quality care and inform strategies for improvement.

This review has limitations that need to be acknowledged. First, the current study findings may not be generalizable to countries with different routine health information systems. Second, while I used a framework to guide the analysis, extraction and classification of indicators were done by a single rater. Third, I focused on indicators available in the Rwanda HMIS reporting forms in this study, which excluded other available routine health information systems in Rwanda, including insurance claims databases, electronic health records, and the community health database. Studies adding these data systems would complement this analysis and help inform future work to create new health system quality indicators that can help identify weaknesses and track the health system performance and ultimately support the use of data in decision making to improve health system quality.

## Conclusion

I found that the Rwanda HMIS captures some processes of care quality indicators such as competent care and system and quality impact indicators such as health outcomes (including mortality, stillbirth and morbidity). However, there were gaps in the measurement of relevant quality impacts such as confidence in health systems and economic benefit, and processes such as user experience. Information about competent care and systems was rarely available outside MNCH. The current Rwanda health information system would benefit from capturing additional healthcare quality metrics, to allow the effective tracking of performance of the health systems and to identify new potential efficiencies. Judicious selection of quality indicators should be central to revising / adding new indicators in order to capture what matters most to patients without overburdening the system with data collection.

### What is already know on this topic

- Effective tracking of the health system performance is vital to identify new potential efficiencies to optimize health system impacts.
- Routine health information systems can provide timely and longitudinal population-based information, making it possible to effectively track the performance of health systems to identify new potential efficiencies in a timely manner.
- Understanding the extent to which these systems can be used to measure quality is imperative given evidence suggests that achieving Sustainable Development Goal 3 will require high-quality health systems in low- and middle-income countries including Rwanda.

### What this study adds

- The Rwanda routine health information systems capture some processes of care quality indicators such as competent care and system and quality impact indicators such as health outcomes (including mortality and morbidity).
- Gaps in the measurement of relevant quality impacts such as confidence in health systems and economic benefit, and processes such as user experience. Information about competent care and systems was rarely available outside maternal, newborn and child health.
- The current routine health information systems in Rwanda would benefit from capturing additional healthcare quality metrics especially for noncommunicable diseases, to allow the effective tracking of the health system performance and to identify new potential efficiencies to maximize the impact of the Rwandan health system.

## Data Availability

All data produced in the present work are contained in the manuscript

## Authors’ contributions

The author conceived and designed the study, performed the analysis and wrote the manuscript.

## Acknowledgements

Dr. Hategeka received support through a Vanier Canada Graduate Scholarship and a Banting Postdoctoral Fellowship. I am grateful to Drs. Law, Lynd and Kenyon for reviewing an initial version of this manuscript.

