## Supplementary material for "Measuring health system quality with routine health information systems in Rwanda": Hategeka Supplemental materials 2024.docx

Celestin Hategeka, MD PhD

**SUPPLEMENTAL MATERIALS**

Appendix Table 1. Processes of care quality indicators from the Rwanda HMIS mapped into the high-quality health system framework

| **Processes of care** | **Indicators or Measures** | **MNCH services** | **Level of care** | **Numerator** | **Denominator** |
| --- | --- | --- | --- | --- | --- |
| **Competent care** |  |  |  |  |  |
| Assessment | Tested for HIV (and received results) during ANC | ANC | Primary | Number of pregnant women who were tested for HIV during ANC visit and who received results | ANC new registrations |
| Assessment | Syphilis testing | ANC | Primary | Number of pregnant women who were tested for syphilis during ANC visit | ANC new registrations |
| Assessment | ANC anemia screening | ANC | Primary | Number of pregnant women who were screened for anemia during ANC visit | ANC new registrations |
| Assessment | Malnutrition screening (MUAC) | ANC | Primary | Number of pregnant women who were screened for malnutrition using MUAC during ANC visit | ANC new registrations |
| Assessment | UTI screening at all ANC visits | ANC | Primary | Number of pregnant women who were screened for UTI during ANC visit | AN new registrations |
| Counseling | Pregnant women counselled during ANC visit and selected PPFP method | ANC | Primary | Number of pregnant women who were counselled and selected PPFP during ANC visit | ANC new registrations |
| Prevention / Treatment | Antibiotics for PPROM | Labor and delivery care | Primary, secondary, tertiary | Women consulted with Preterm Premature Ruptured Membranes (PPROM) who received prophylactic antibiotics | Women consulted with Preterm Premature Ruptured Membranes (PPROM) |
| Prevention / Treatment | Oxytocin for AMTSL | Labor and delivery care | Primary, secondary, tertiary | Women who received oxytocin IM immediately after birth for active management of third stage of labor (AMTSL) | Number of deliveries in a health facility |
| Prevention / Treatment | Corticosteroid for risk of premature delivery | Labor and delivery care | Primary, secondary, tertiary | Mother who received Corticosteroid in management of risk of premature delivery | Woman consulted for risk of premature delivery |
| Treatment | Obstetrical fistula conservative treatment with Foley catheter | Labor and delivery care | Primary, secondary, tertiary | Number of women suspected to develop obstetrical fistula (during labor) who received conservative treatment with Foley catheter | Number of women suspected to develop obstetrical fistula during labor |
| Treatment | Successful neonatal resuscitation | Labor and delivery care | Primary, secondary, tertiary | Number of live newborns who didn’t cry/breathe at birth and were resuscitated successfully (cry/breath within 5 minutes, APGAR score >5 at 5min | Number of live newborns who didn’t cry at birth and for whom resuscitation was performed using ambu bag |
| **Competent systems** |  |  |  |  |  |
| Prevention / Treatment | Iron and Folic acid supplementation among women who attended ANC | ANC | Primary | Women who were given full course of iron and folic acid supplements | ANC new registrations |
| Timely action | Breastfeeding within 1 hour | Immediate newborn care | Primary, secondary, tertiary | Number of newborns breastfed within 1 hour of delivery in a health facility | Number of live births in a health facility |
| Timely action | Newborns placed skin to skin after birth for at least for one hour | Immediate newborn care | Primary, secondary, tertiary | Number of newborns placed skin to skin after birth for at least for one hour in a health facility | Number of live births in a health facility |
| Timely action | PNC1 mothers | PNC | Primary, secondary, tertiary | Newborn PNC1 visits within 24 hours of birth | Number of live births in a health facility |
| Timely action | PNC1 newborns | PNC | Primary, secondary, tertiary | Mother PNC1 visits within 24 hours of birth | Number of deliveries in a health facility |

ANC, antenatal care; PNC, postnatal care; AMTSL, active management of third stage of labor; UTI, urinary tract infection; MUAC, Mid-Upper Arm Circumference; HIV, human immunodeficiency virus; PPFP, postpartum family planning; APGAR, Appearance, Pulse, Grimace, Activity, Respiration; PPROM, Preterm Premature Ruptured Membranes.

Appendix Table 2. Quality impact indicators from the Rwanda HMIS mapped into the high-quality health system framework

| **Health outcomes** | **Indicators or Measures** | **Numerator** | **Denominator** |
| --- | --- | --- | --- |
| Mortality | Obstetrical complication case fatality rate | Women who died of obstetrical complications | Women admitted to a health facility for obstetrical complications in a health facility |
| Mortality | Early maternal mortality ratio | Women who died during labor, delivery, and within 24 hours postpartum | Number of live births in a health facility |
| Stillbirth | Stillbirth rate | All stillbirths (antepartum stillbirths + intrapartum stillbirths) | Births (live births + stillbirths) in a health facility |
| Stillbirth | Intrapartum stillbirth rate | Stillbirths who are fresh and without any signs of maceration | Births in a health facility |
| Stillbirth | Antepartum stillbirth rate | Stillbirths who are macerated | Births in a health facility |
| Mortality | 30-minute neonatal mortality rate | Number of newborn deaths within 30 minutes of birth | Live births in a health facility. |
| Mortality | Neonatal hospital mortality rate | Number of deaths in neonatology department (Newborn deaths within 28 days) | Number of newborns admitted to a health facility (new admission and present at the beginning of the month). |
| Mortality | Pediatric hospital mortality rate | Number of deaths in pediatric department (post-neonatal period) | Number of children admitted to pediatric department in a health facility (post-neonatal period) |
| Mortality | Under-five hospital mortality rate | Number of deaths among children younger than five years in a health facility | Number of admissions among children younger than five years in a health facility |
| Mortality | Obstetrics and gynecology mortality rate | Number of deaths in obstetrics and gynecology department in a health facility | Number of obstetrics and gynecology admissions in a health facility |
| Mortality | Internal medicine mortality rate | Number of deaths in internal medicine department in a health facility | Number of internal medicine admissions in a health facility |
| Mortality | Surgical mortality rate | Number of deaths in surgery department in a health facility | Number of surgery admissions in a health facility |
| Morbidity | Neonatal asphyxia incidence | Number of neonatal asphyxia hospital admission | Live births |
| Morbidity | Birth trauma to newborns | Number of newborns with birth trauma | Live births |
| Morbidity | Uterine rupture | Number of uterine ruptures | hospital deliveries |
| Morbidity | Perineal laceration | Number of perineal lacerations | Hospital deliveries |
| Morbidity | Post C/S surgical site infection | Women with infections at the C/S surgical site | Women who delivered by caesarean section |
| Morbidity | PPH incidence | Women with PPH | Number of facility deliveries |
| Morbidity | Obstetrical fistula | Women with obstetrical fistula | Number of facility deliveries |
| Morbidity | Hysterectomy for treatment of PPH | Women who had hysterectomy for treatment of PPH | Number of facility deliveries |
| Morbidity | Postpartum infection rate | Women with postpartum infections | Number of facility deliveries |

Appendix Table 3. STROBE Statement—Checklist of items that should be included in reports of ***cross-sectional studies***

|  | Item No | Recommendation | Page No |
| --- | --- | --- | --- |
| **Title and abstract** | 1 | (*a*) Indicate the study’s design with a commonly used term in the title or the abstract | 2 |
|  |  | (*b*) Provide in the abstract an informative and balanced summary of what was done and what was found | 2 & 3 |
| Introduction | | | |
| Background/rationale | 2 | Explain the scientific background and rationale for the investigation being reported | 4 & 5 |
| Objectives | 3 | State specific objectives, including any prespecified hypotheses | 6 |
| Methods | | | |
| Study design | 4 | Present key elements of study design early in the paper | 6 |
| Setting | 5 | Describe the setting, locations, and relevant dates, including periods of recruitment, exposure, follow-up, and data collection | 6 |
| Participants | 6 | (*a*) Give the eligibility criteria, and the sources and methods of selection of participants | NA |
| Variables | 7 | Clearly define all outcomes, exposures, predictors, potential confounders, and effect modifiers. Give diagnostic criteria, if applicable | 6-7 |
| Data sources/ measurement | 8* | For each variable of interest, give sources of data and details of methods of assessment (measurement). Describe comparability of assessment methods if there is more than one group | *6-7, Appendix tables 1 & 2* |
| Bias | 9 | Describe any efforts to address potential sources of bias | NA |
| Study size | 10 | Explain how the study size was arrived at | NA |
| Quantitative variables | 11 | Explain how quantitative variables were handled in the analyses. If applicable, describe which groupings were chosen and why | NA |
| Statistical methods | 12 | (*a*) Describe all statistical methods, including those used to control for confounding | 7-8 |
|  |  | (*b*) Describe any methods used to examine subgroups and interactions | NA |
|  |  | (*c*) Explain how missing data were addressed | NA |
|  |  | (*d*) If applicable, describe analytical methods taking account of sampling strategy | NA |
|  |  | (*e*) Describe any sensitivity analyses | NA |
| Results | | | |
| Participants | 13* | (a) Report numbers of individuals at each stage of study—eg numbers potentially eligible, examined for eligibility, confirmed eligible, included in the study, completing follow-up, and analysed | NA |
|  |  | (b) Give reasons for non-participation at each stage | NA |
|  |  | (c) Consider use of a flow diagram | NA |
| Descriptive data | 14* | (a) Give characteristics of study participants (eg demographic, clinical, social) and information on exposures and potential confounders | 8-10 |
|  |  | (b) Indicate number of participants with missing data for each variable of interest | NA |
| Outcome data | 15* | Report numbers of outcome events or summary measures | 8-10 |
| Main results | 16 | (*a*) Give unadjusted estimates and, if applicable, confounder-adjusted estimates and their precision (eg, 95% confidence interval). Make clear which confounders were adjusted for and why they were included | NA |
|  |  | (*b*) Report category boundaries when continuous variables were categorized | NA |
|  |  | (*c*) If relevant, consider translating estimates of relative risk into absolute risk for a meaningful time period | NA |
| Other analyses | 17 | Report other analyses done—eg analyses of subgroups and interactions, and sensitivity analyses | NA |
| Discussion | | | |
| Key results | 18 | Summarise key results with reference to study objectives | 10 |
| Limitations | 19 | Discuss limitations of the study, taking into account sources of potential bias or imprecision. Discuss both direction and magnitude of any potential bias | 12-13 |
| Interpretation | 20 | Give a cautious overall interpretation of results considering objectives, limitations, multiplicity of analyses, results from similar studies, and other relevant evidence | 10-12 |
| Generalisability | 21 | Discuss the generalisability (external validity) of the study results | 12 |
| Other information | | | |
| Funding | 22 | Give the source of funding and the role of the funders for the present study and, if applicable, for the original study on which the present article is based | NA |

*Give information separately for exposed and unexposed groups.

**Note:** An Explanation and Elaboration article discusses each checklist item and gives methodological background and published examples of transparent reporting. The STROBE checklist is best used in conjunction with this article (freely available on the Web sites of PLoS Medicine at http://www.plosmedicine.org/, Annals of Internal Medicine at http://www.annals.org/, and Epidemiology at http://www.epidem.com/). Information on the STROBE Initiative is available at www.strobe-statement.org.
